# Two decades of molecular surveillance in Senegal reveal changes in known drug resistance mutations associated with historical drug use and seasonal malaria chemoprevention

**DOI:** 10.1101/2023.04.24.23288820

**Authors:** Yaye Die Ndiaye, Wesley Wong, Julie Thwing, Stephen S Schaffner, Abdoulaye Tine, Mamadou Alpha Diallo, Awa Deme, Mouhammad Sy, Amy K Bei, Alphonse B Thiaw, Rachel Daniels, Tolla Ndiaye, Amy Gaye, Ibrahima Mbaye Ndiaye, Mariama Toure, Nogaye Gadiaga, Aita Sene, Djiby Sow, Mamane N. Garba, Mamadou Samba Yade, Baba Dieye, Khadim Diongue, Daba Zoumarou, Aliou Ndiaye, Jules Gomis, Fatou Ba Fall, Medoune Ndiop, Ibrahima Diallo, Doudou Sene, Bronwyn Macinnis, Mame Cheikh Seck, Mouhamadou Ndiaye, Aida S. Badiane, Daniel L. Hartl, Sarah K. Volkman, Dyann F. Wirth, Daouda Ndiaye

## Abstract

Drug resistance in *Plasmodium falciparum* is a major threat to malaria control efforts. We analyzed data from two decades (2000-2020) of continuous molecular surveillance of *P. falciparum* parasite strains in Senegal to determine how historical changes in drug administration policy may have affected parasite evolution. We profiled several known drug resistance markers and their surrounding haplotypes using a combination of single nucleotide polymorphism (SNP) molecular surveillance and whole-genome sequence (WGS) based population genomics. We observed rapid changes in drug resistance markers associated with the withdrawal of chloroquine and introduction of sulfadoxine-pyrimethamine in 2003. We also observed a rapid increase in *Pfcrt* K76T and decline in *Pfdhps* A437G starting in 2014, which we hypothesize may reflect changes in resistance or fitness caused by seasonal malaria chemoprevention (SMC). Parasite populations evolve rapidly in response to drug use, and SMC preventive efficacy should be closely monitored.

## INTRODUCTION

The World Health Organization (WHO) estimated 247 million malaria cases and 619,000 deaths in 2021^1^. Children under 5 years of age are the most vulnerable to malaria, accounting for 80% of deaths worldwide; 96% of malaria cases and deaths in 2021 were in the African region^1^. Increased funding for malaria control has led to an estimated 30% reduction of malaria mortality since 2000, using a combination of vector control and drug-based interventions^2^. However, the development of parasite resistance to antimalarials threatens to undermine control and elimination efforts and poses a significant threat to public health.

Therapeutic efficacy studies (TES) are the gold standard for evaluating clinical and therapeutic drug efficacy^3,4^. TES are prospective evaluations of patients’ clinical and parasitological responses to treatment as assessed 28 or 42 days after treatment. However, TES are resource intensive and can be challenging to implement where transmission is low^5^. Furthermore, antimalarial drugs may also be used as chemoprophylaxis to prevent infection. WHO-recommended chemopreventive strategies include intermittent preventive treatment for pregnant women (IPTp) and infants (IPTi), seasonal malaria chemoprevention (SMC), and mass drug administration (MDA)^1,6^. As with TES, chemopreventive efficacy studies can be challenging to implement and require significant planning and longitudinal monitoring^7^.

Parasite drug resistance is a major factor that influences therapeutic and chemopreventive antimalarial efficacy. Molecular surveillance of genetic markers could be used to detect emerging drug resistance or changes in parasite fitness ^8–13^ that could undermine therapeutic or chemopreventive efficacy. Molecular surveys of drug resistance commonly focus on assessing the frequency of single nucleotide polymorphisms (SNPs) or copy number variations that have been associated with drug resistance in laboratory settings^1,13^. Scanning the genomic regions and haplotypes surrounding these mutations for evidence of hard or soft selective sweeps also provides insight into the origins of these mutations and provides additional evidence of drug selection at these sites^13,14^. Selective sweeps occur when mutations become fixed or are eliminated so rapidly that it causes a reduction in the variation at nearby nucleotide positions^13,14^. Hard selective sweeps indicate selection from a single genomic background, while soft selective sweeps indicate selection from multiple backgrounds or pre-existing standing variation.

In this study, we examined a two-decade collection of *P. falciparum* samples collected from febrile individuals in Senegal to assess how historical changes in drug use has affected the parasite population. Based on the historical antimalarial policy in Senegal, we focused our analyses on SNPs associated with resistance to chloroquine (CQ), amodiaquine (AQ), sulfadoxine-pyrimethamine (SP), and artemisinin (ART). We were also interested in using molecular surveillance to look for signs of emerging drug resistance or changes in parasite fitness that could signal a reduction in IPTp or SMC chemopreventive efficacy in Senegal. In Senegal, IPTp is administered as a dose of SP to pregnant women during antenatal care visits and is spaced at least 30 days apart. A single cycle of SMC consists of a dose of SP + AQ on the first day, followed by doses of AQ on days 2 and 3. In Senegal, children 3 months to 10 years are targeted, with three to five monthly cycles during the transmission season, depending on the length of the transmission season in their region.

## RESULTS

### Study design and SNP-based molecular surveillance sampling

*P. falciparum* samples from febrile patients collected between 2000-2020 were genotyped. A total of 3,284 samples were collected from six regions of Senegal: Pikine, Thiès, Kedougou, Diourbel, Kaolack, and Kolda (**Figure 1A**). Kedougou and Kolda are high transmission regions in southeast Senegal. In 2021, reported annual incidence was 536.5 cases per 1000 in Kedougou and 214.5 cases per 1000 in Kolda^15^. SMC has been implemented in Kedougou and Kolda since 2014. Kaolack (10.9 cases per 1000) and Diourbel (19.4 cases per 1000) are intermediate transmission regions in central Senegal, and started SMC in 2019. Thiès (2.8 cases per 1000) and Pikine (4.9 cases per 1000) are low transmission sites in western Senegal. SMC is not implemented in Thiès or Pikine.

Our sample collection spans several important changes in official drug use policy (**Fig 1B**). These changes include the withdrawal of CQ and the introductions of SP, AQ, and artemisinin-based combination therapies (ACTs). Based on this drug policy history, we examined several SNP-based drug resistance markers in *Pfcrt*, *Pfdhfr*, *Pfdhps*, *Pfmdr1*, and *Pfklech13* (**Table 1**). Because resistance to pyrimethamine involves multiple mutations in *Pfdhfr*, we examined the frequency of *Pfdhfr* triple mutant CRN (mutant at N51C, C59R, and S108N)^16^ and “quadruple” mutants (*Pfdhfr* triple mutant CRN + *Pfdhps* A437G). Likewise, we examined the frequency of the *Pfmdr1* NFD (N86Y, Y184F, D1246Y) haplotype because the haplotype is associated with resistance against multiple drugs^17,18^.

**Fig 1:**
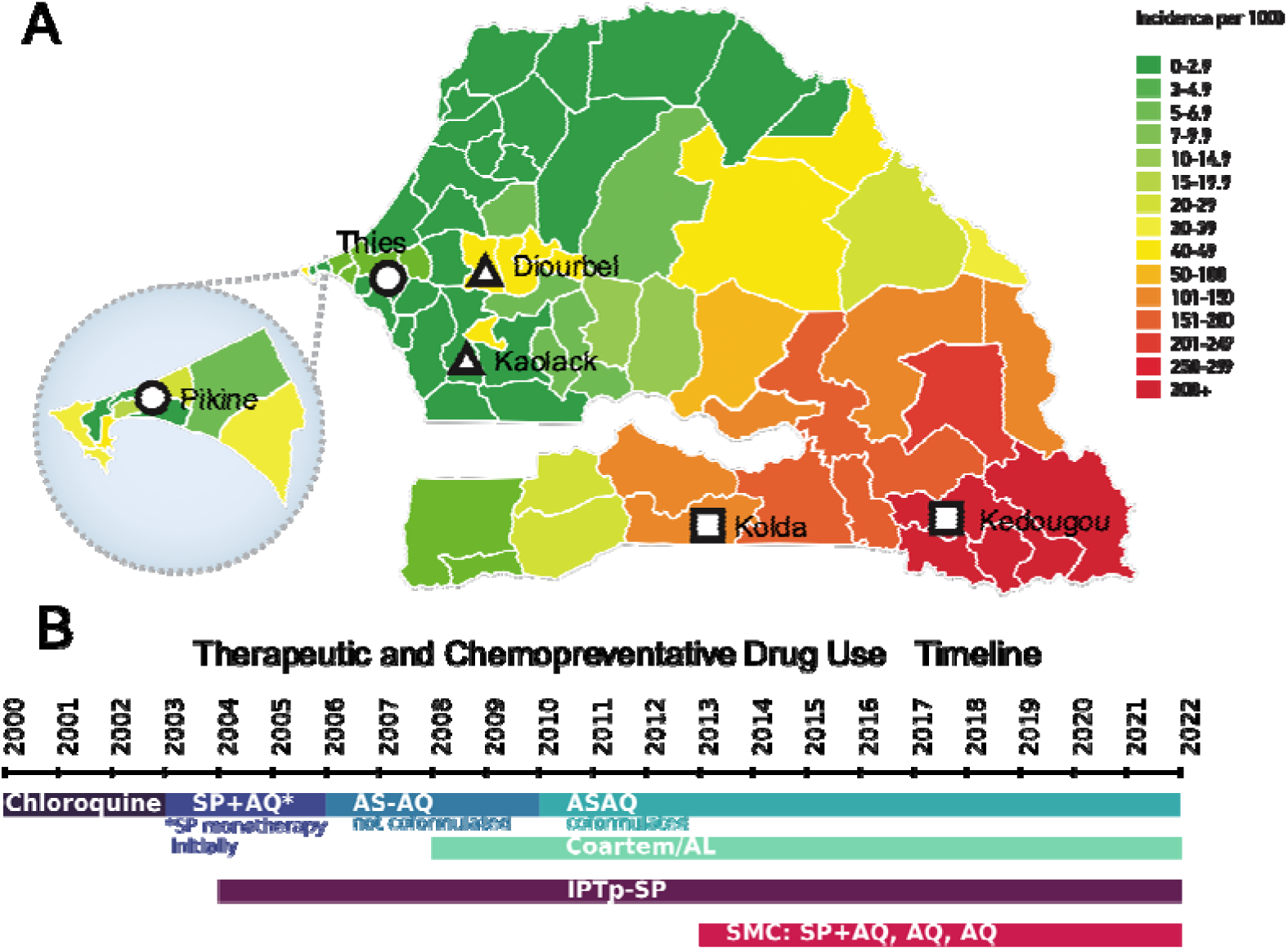
Study design and Senegal drug history. **A**) Map of Senegal highlighting the different study locations and their corresponding transmission levels in 2019. Each region is colored by transmission intensity, with red indicating high transmission and green indicating low. Squares denote sampled regions that started SMC in 2014, triangles those that started in 2019, and circles those that have not implemented SMC. **B**) Therapeutic and chemopreventive drug use in Senegal. Therapeutic drug use in *blue/green*, chemopreventive drug use in *red/purple*. SP = Sulfadoxine-Pyrimethamine, AQ = amodiaquine, ASAQ = Artesenuate/Amodiaquine, Coartem/AL = Coformulated Artemether Lumefantrine, IPTp-SP = Intermittent preventative therapy in pregnant women using SP, SMC = Seasonal malaria chemoprevention.

**Table 1.** Table of the molecular markers examined and their corresponding drug resistances.

| Gene | SNPs assayed | Expected Drug Resistance |
| --- | --- | --- |
| Pfcrt | K76T | Chloroquine, Amodiaquine |
| PfKelch13 | A578S, C580Y | Artemisinin |
| Pfdhps | A437G, K540E, A581G, A613T/S | Sulfadoxine |
| Pfdhfr | N51C, C59R, S108N | Pyrimethamine |
| Pfmdr1 | N86Y, Y184F, D1246Y | Multiple, including Amodiaquine, Mefloquine, Chloroquine, Artemether-Lumefantrine |

Our sample collection varied throughout time and space (**Fig S1**). The 444 samples collected between 2000 and 2005 came exclusively from Pikine, the 800 samples collected between 2006 and 2014 came from Thiès, and the 340 samples collected between 2015 and 2017 came from Kedougou. Between 2017 and 2020, 1700 samples were collected from Thiès, Kaolack (starting in 2020), Diourbel, Kolda (starting in 2019), and Kedougou.

### Molecular surveillance detects rapid changes in *Pfcrt*, *Pfdhfr*, *Pfdhps*, and *Pfmdr1* mutations over time

The changes in sampling numbers and locations over time presented a unique challenge. We first examined the population frequencies of each SNP in the sample regions with more than three years of continuous sampling: Pikine, Thiès, and Kedougou (**Fig S2-S4**). Pikine and Thies are urban sites with low transmission that have not implemented SMC while Kedougou is a rural site with high transmission that has been implementing SMC since 2014.

We noted that the data from Pikine and Thiès (which are approximately 30 miles apart) prior to 2014 appeared to be part of a continuous time series and that changes in Thiès and Kedougou after 2014 followed similar trends. To summarize the molecular surveillance data and identify broad changes in mutation frequency, we used a binomial generalized additive model (GAM) to identify and highlight Senegal-wide trends in mutation frequency (**Methods**, **Fig 2**).

**Fig 2.**
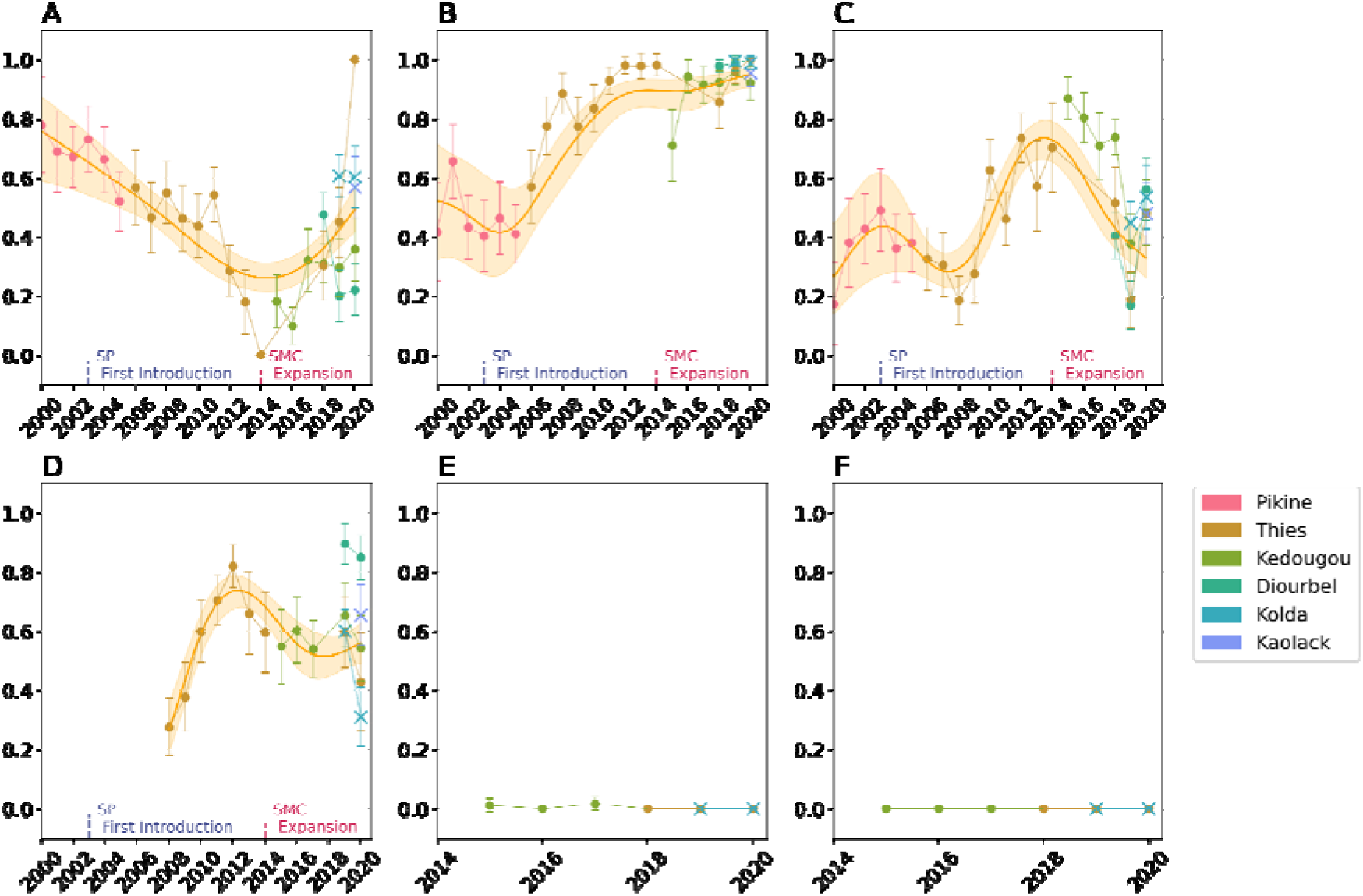
SNP-based Molecular Surveillance Results. Frequencies for **A**) *Pfcrt* K76T, **B**) *Pfdhfr* triple mutant (N51C, C59R, S108N), **C**) *Pfdhps* A437G, **D**) *Pfmdr1* N_86_F_184_D_1246_ haplotype (N86Y, Y184F, D1246Y), **E**) *Pfkelch13* A578S, and **F**) *Pfkelch13* C580Y. The scatterplots show the observed frequencies and their 95% binomial confidence interval. Model predictions from a calibrated generalized additive model and the 95% confidence intervals are shown in orange. The model was calibrated with data from Pikine, Thiès, Diourbel, and Kedougou (denoted with circles). The data from Kolda and Kaolack (denoted with X) were not used for model calibration. Model predictions were not generated for the *Pfkelch13* mutations due to their complete or near-complete absence in the data.

#### Increase in Pfcrt K76T mutation frequency after 2014

As expected, due to a fitness cost associated with *Pfcrt K76T,* we observed a decline in *Pfcrt K76T* mutation frequency between 2000 and 2014 following the withdrawal of CQ in 2003. Unexpectedly, this was followed by an increase in frequency after 2014. In 2000, our model estimated a Senegal-wide mutation frequency of 0.76 (95% CI 0.59 - 0.87), which fell to 0.26 (95% CI0.21 - 0.31) in 2014. However, the frequency began rising after 2014, and the model estimated the frequency to be 0.49 (95% CI 0.41 - 0.57) in 2020. The adjusted *R*-squared for the model was 0.466 and the deviance explained 54.2%.

When examined separately, there was a statistically significant increase in *Pfcrt* K76T in both Thiès (p-value < 0.00001) and Kedougou (p-value = 9.6e-5) after 2014. When examining the sites not included in the Senegal-wide GAM (Kolda, and Kaolack), the frequency of *Pfcrt K76T* were consistent with those predicted by the GAM. However, the frequency of *Pfcrt K76T* in Diourbel trended in the opposite direction, declining from 0.45 (95% CI 0.55 - 0.40) to 0.20 (95% CI 0.11 - 0.29) and 0.22 (95% CI 0.13 - 0.31) in 2018, 2019, and 2020, respectively.

#### Changes in Pfdhfr triple mutants following the withdrawal of CQ and introduction of SP

We expected an increase in *Pfdhfr CRN* triple mutants due to pyrimethamine exposure from either SP therapy beginning in 2003 or IPTp chemoprevention beginning in 2004. Our data show a sharp rise in the frequency of *Pfdhfr* triple mutants starting in 2003, coinciding with the replacement of CQ with SP as the first-line antimalarial treatment in Senegal (**Fig 2B**). The increase in the *Pfdhfr* triple mutant corresponded to a decrease in *Pfdhfr* triple sensitives (*N51N, C59C, S108S*) (**Fig S5A**) and we detected only a few parasites with a mix of *Pfdhfr N51N, C59C, S108S* status. In 2003, the GAM predicted that the frequency of triple mutants in Senegal was 0.42 (95% CI 0.27 - 0.59). By 2020, the predicted frequency was 0.95 (95% CI 0.90 - 0.97). The adjusted R-squared and deviance explained by the model were 0.881 and 89.4%.

#### Rise and fall of Pfdhps A437G before and after SMC expansion in 2014

We expected mutations in *Pfdhps* to increase due to sulfadoxine exposure from either SP therapy or IPTp chemoprevention or SMC. *Pfdhps* K540E, A581G, A613T/S were rare (< 5%) or undetected by our sampling and only *Pfdhps* A437G was detected at high frequencies (**Fig 2C**, **Fig S2-S4**). Unlike with the *Pfdhfr* triple mutants, the *Pfdhps A437G* allele trajectory and “quadruple” mutant haplotype trajectory changed directions multiple times (**Fig S5B**). Our data revealed several inflection points (at 2003, 2008, and 2014) where the trajectory of *Pfdhps* prevalence changed.

Between 2000 and 2003, we observed an increase in *Pfdhps* A437G from the GAM-predicted frequency of 0.26 (95% CI 0.14 - 0.44) in 2000 to 0.47 (95% CI 0.19 - 0.77) in 2003. After 2003, *Pfdhps* A437G decreased until 2008, when its predicted frequency was 0.17 (95% CI 0.11 - 0.27). Between 2008 and 2014, the frequency of *Pfdhps A437G* rose until 2014, when its predicted mutation frequency was 0.72 (95% CI 0.58 - 0.83). After 2014, the frequency of *Pfdhps* A437G declined, and the predicted mutation frequency in 2020 was 0.33 (95% CI 0.25 - 0.40). Overall, the adjusted *R*-squared and deviance explained by the model were 0.554 and 69.1%, respectively.

#### Molecular surveillance detects changes in Pfmdr1 NFD haplotype over time

Expectations for the *Pfmdr1* NFD were less certain as mutations in *Pfdmr1* have been associated with resistance against multiple drugs. Between 2008 and 2012, we observed a rapid increase in the *Pfmdr1* NFD (N86Y, Y184F, D1246Y) haplotype (**Fig 2D**). The corresponding model-predicted frequencies were 0.27 (95% CI 0.20 - 0.37) in 2004 and 0.74 (95% CI 0.57 - 0.79) in 2012. After 2012, the predicted frequency of the *Pfmdr1* NFD haplotype declined to 0.56 (95% CI 0.48 - 0.63) in 2016, where it remained relatively stable until 2020 [0.56 (95% CI 0.49 - 0.63)]. The adjusted *R*-squared and deviance explained by the model were 0.817 and 87.2%, respectively.

#### Infrequent detection of Pfkelch13 A578S and absence of Pfkelch13 C580Y in Senegal

*Pfkelch13* C580Y was not detected in our samples, while *Pfkelch13 A578S* was present but infrequently detected (**Fig 2E-F**). The *Pfkelch13* A578S mutation was observed in one of the 89 genotyped samples collected from Kedougou in 2015 and twice in the 123 genotyped samples collected from Kedougou in 2017, but in no other year. The *Pfkelch13* C580Y mutation was not detected in any of our genotyped samples.

#### Genomic haplotype analyses reveal differences in selection acting on Pfcrt, Pfdhps, and Pfdhfr

To determine whether the changes in *Pfcrt*, *Pfdhps*, and *Pfdhfr* resulted from drug-mediated selection, we examined the genomic haplotypes surrounding these genes in a set of 231 whole genome sequences collected from Thiès and Kedougou between 2006 and 2019 (**Fig S6**). Our primary goal was to determine whether *Pfcrt* K76T, *Pfdhps* A437G, and the *Pfdhfr* triple mutant CRN showed evidence of a rapid selective sweep indicative of strong drug pressure by examining the haplotype diversity in the surrounding genomic regions. *Pfmdr1* was not examined because of its strong potential for copy number variation.

For *Pfcrt* K76T, we found strong evidence of a selective sweep. The haplotype structure surrounding *Pfcrt* K76T was far less diverse than that surrounding the ancestral *Pfcrt* K76 (**Fig 3A-B**). *Pfcrt* K76 haplotypes rapidly diversified within the first 20kb. In contrast, *Pfcrt* K76T was surrounded by a dominant extended haplotype to its left (upstream) and two major extended haplotypes to its right (downstream). These dominant haplotypes extended out nearly 50 kb away from the *Pfcrt* K76T locus and included parasites collected before and after 2014. The extended haplotype homozygosity (EHH) for *Pfcrt* K76T was elevated relative to *Pfcrt* K76 (**Fig 3C**).

**Fig 3.**
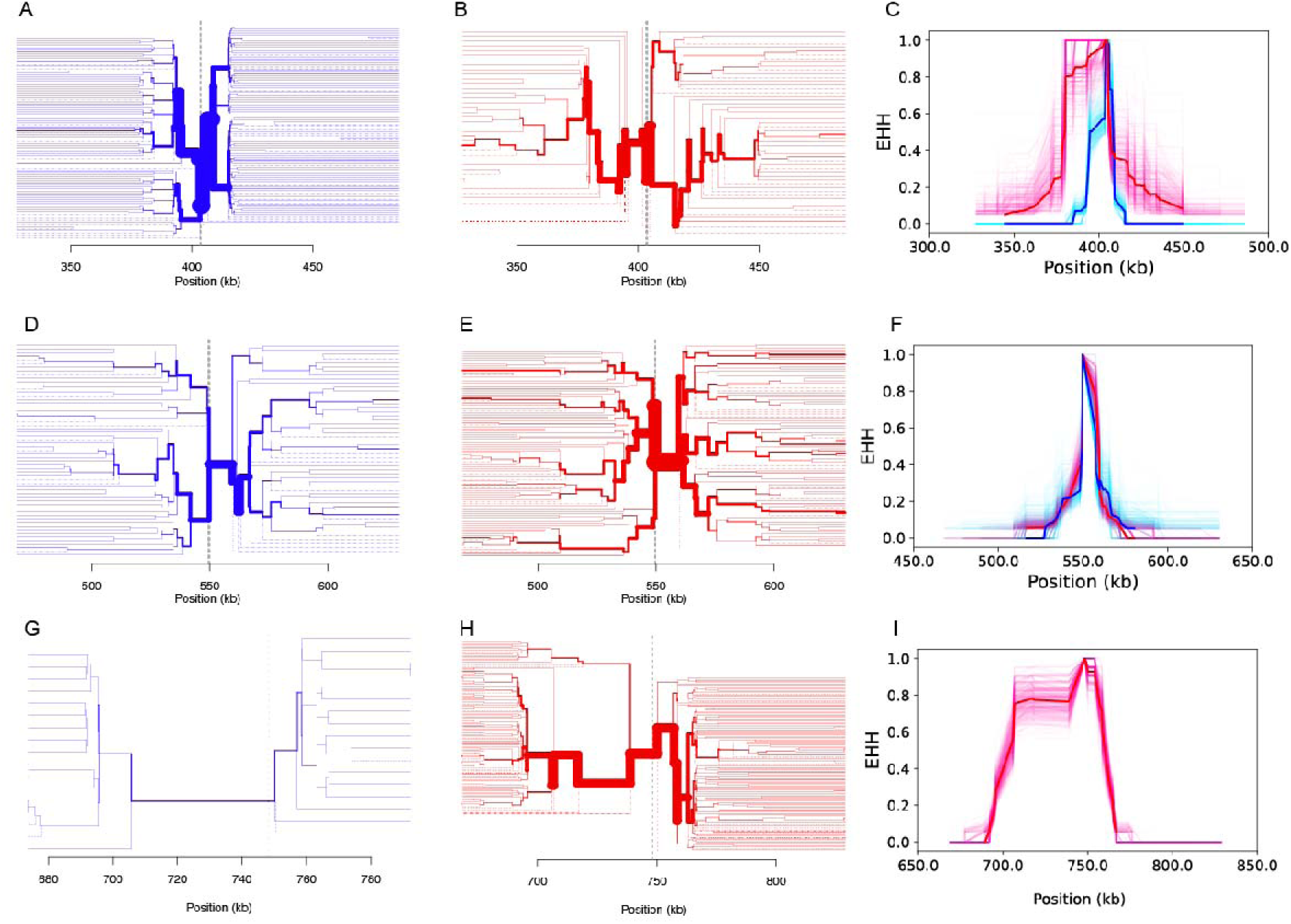
Evidence of selection at *Pfcrt, Pfdhps, and Pfdhfr*. Extended haplotype plots for the **A-C**) *Pfcrt* K76T locus, **D-F**) *Pfdhps* A437G locus, and **G**) the *Pfdhfr* triple mutant. Red is used for the resistance allele (*Pfcrt* K76T*, Pfdhps* A437G, and *Pfdhfr* triple mutant) while blue is used for the sensitive allele (*Pfcrt* K76*, Pfdhps* A437*, Pfdhfr* triple sensitive). Column 1 (**A, D, G**) used samples collected before and after the implementation of SMC. Column 2 (**B, E**) used samples collected before SMC (before 2014), and Column 3 (**C**, **F**) used samples collected after SMC (after 2014). The solid red and blue lines are the EHH estimates obtained from using all available samples in the category. The lighter pink and blue traces are bootstrapped EHH estimates obtained by randomly downsampling fifty samples. For **G-I**, samples with mixed *Pfdhfr* genotypes were excluded.

For *Pfdhps*, we surprisingly found little evidence of a hard selective sweep (selection of a single or small number of haplotypes). Instead, the haplotype structure surrounding the sensitive and resistant alleles were highly diverse and there was no significant elevation in EHH (**Fig 3D-F**). Multiple long-range haplotypes extended outwards from either *Pfdhps* A437G or *Pfdhps* A437A for 30-50 kb. Combined with the changes in allele frequency observed in the molecular surveillance, we wanted to determine whether *Pfdhps* A437G showed evidence of being selected for but across multiple competing backgrounds (a “soft” sweep)^19^. To test this, we adapted two existing statistical measures of selection, H12 and H2/H1. H12 is the expected haplotype homozygosity, treating the two most common haplotypes as if it were a single haplotype, and H2/H1, the ratio of the second and the first most frequent haplotype (**Fig S7**)^20^. Elevated H12 and a low H2/H1 ratio indicates a hard selective sweep while elevated H12 and a high H2/H1 ratio suggests a soft selective sweep (**Methods**). We found that, compared to the rest of chromosome 8, both H12 and the H2/H1 ratio around *Pfdhps* were elevated (**Fig S7A**), consistent with a soft sweep. Conversely, H12 around *Pfcrt* was high but H2/H1 was low, indicating a harder selective sweep (**Fig S7B**).

We detected a strong extended haplotype structure surrounding the *Pfdhfr* triple mutant. Most *Pfdhfr* triple mutants shared the same dominant haplotype (**Figure 3 G-I**). This haplotype extended out more than 50 kb to the left of the *Pfdhfr* gene and 20 kb to the right. The EHH of the *Pfdhfr* was likely due to a hard selective sweep, similar to that detected for *Pfcrt* K76T. However, we lacked the sensitivity to accurately estimate the EHH surrounding the *Pfdhfr* triple sensitive genotype. Most samples with usable *Pfdhfr* sequences were collected after 2010 (**Fig S5C**), after the rapid increase in the *Pfdhfr* triple mutant identified by our SNP-based molecular surveillance (**Fig 2C**). *Pfdhfr* triple mutants comprised 0.83 (95% CI 0.77 - 0.89) of our whole genome sequenced samples; 0.07 (95% CI 0.03 - 0.11) were mixed mutants, and only 0.10 (95% CI 0.05 - 0.14) of our samples were *Pfdhfr* triple sensitive parasites.

#### Haplotype analyses reveal temporal changes in selection at Pfcrt K76T before and after 2014

For *Pfcrt* K76T, we also estimated the EHH before and after SMC to determine whether SMC could be driving the selective sweep (**Fig 4**). Prior to 2014, the EHH directly proximal to the *Pfcrt* K76T resistance mutation was similar to that observed surrounding the sensitive *Pfcrt* K76. However, we detected elevated EHH 10 kb upstream and downstream of the *Pfcrt* K76T locus, which is likely a legacy of the historical selective sweep that occurred prior to CQ withdrawal in 2003 (**Fig 4B**)^21^. After the introduction of SMC in 2014, the EHH surrounding *Pfcrt* K76T was elevated (**Fig 4A, C**) and consistent with the expectations for a new selective pressure acting on the mutation occurring after 2014.

**Fig 4.**
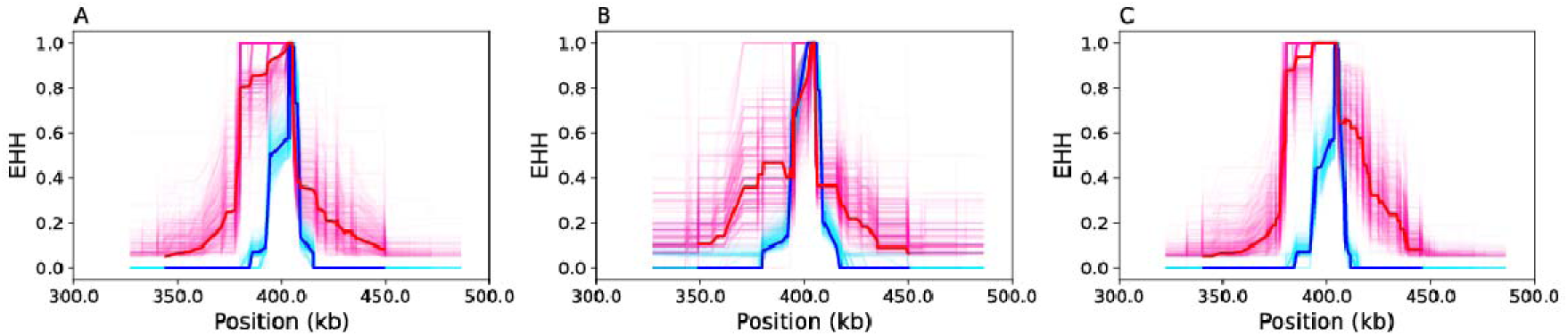
Temporal shifts in selection for *Pfcrt* K76T. Extended haplotype plots for the *Pfcrt* K76T locus using **A)** all collected samples, **B**) samples collected before or during 2014, and samples collected after 2014. The solid red and blue lines are the EHH estimates obtained from using all available samples in the category. The lighter pink and blue traces are bootstrapped EHH estimates obtained by randomly downsampling fifty samples. Note that **Fig 4A** is the same as **Fig 3C**.

## DISCUSSION

Accurate assessments of drug resistance in parasite populations are needed to ensure continued success of drug-based malaria control efforts. The changes in the frequency of *Pfdhfr* triple mutant CRN suggest that pyrimethamine resistance is widespread throughout Senegal. Nearly all parasites carried the *Pfdhfr* triple mutant by 2013. The increase in *Pfdhfr* triple mutant is consistent with pyrimethamine-mediated drug resistance, first from antimalarial SP therapy, and then from chemopreventive IPTp. A similar rise in *Pfdhfr* triple mutant frequency was previously reported by a study carried out in Thiès in 2003 and 2013^22,23^. Our haplotype analysis suggests that SP-mediated pyrimethamine increase in *Pfdhfr* triple mutant occurred rapidly from a single haplotype.

Changes in *Pfcrt K76T* and *Pfdhps A437G* were more complex but coincided with several important changes in either therapeutic or chemopreventive drug use. In 2003, Senegal stopped recommending CQ as the first-line antimalarial therapy due to widespread CQ resistance across the African continent^21,24,25^. Although *Pfcrt K76T* is associated with CQ resistance, it confers a fitness cost in *in vitro* laboratory settings where CQ is absent ^26^. The decline in *Pfcrt* K76T reported in our study is consistent with other molecular surveillance studies that have shown similar declines in frequency after the withdrawal CQ ^27,28^ and similar reductions in EHH (**Figure 4B**) surrounding the *Pfcrt* K76T mutation as parasites begin outcrossing more frequently with CQ sensitive parasites^29,30^.

The increase in *Pfcrt* K76T in 2014 was unexpected but our haplotype analyses suggest that the increase in frequency is due to a new selection event occurring after 2014. These changes coincide with the implementation and expansion of SMC in the high transmission regions of Senegal. We suspect the sudden increase in *Pfcrt* K76T may be being driven by the AQ used in SMC and by the introduction of ASAQ, one of the first line ACTs in use in Senegal. Laboratory based studies have shown that the mutation confers AQ resistance and previous molecular surveillance studies have associated the mutation with AQ resistance ^31,32^. In Nigeria, AQ monotherapy may explain the high frequencies of *Pfcrt* K76T despite the withdrawal of CQ^33^. Despite these findings, recent TES studies in Senegal show that ASAQ remained highly efficacious in 2020^34^. The therapeutic efficacy of ASAQ should be closely monitored, as our data suggest that molecular evidence of AQ resistance is increasing in Senegal. Efforts to ensure that ASAQ is not used for treatment in areas in which SMC is in use are critical.

Likewise, the rapid frequency changes at *Pfdhps* A437G was unexpected and is a reversal of the accumulation of “quadruple” mutant parasites (parasites that are *Pfdhfr* triple mutant and *Pfdhps* A437G) observed in African countries where SP is administered^35^. These results suggest that Senegal parasites may be more sensitive to sulfadoxine relative to the quadruple mutant parasites observed in other African countries. It is unclear what is driving this decrease in *Pfdhps A437G*, but we hypothesize that it is related to an AQ-induced fitness cost. This hypothesis is based primarily on the decline in *Pfdhps* A437G shortly after the implementation of SMC in 2014 but could also explain the decline in *Pfdhps* A437G frequency between 2003 and 2008 when SP-AQ and ASAQ were used for antimalarial therapy. The increase between 2008 and 2014 could be because antimalarial therapies have increasingly relied on Artemether Lumefantrine (Coartem/AL) after its introduction in 2008^36^.

Determining whether the changes in *Pfcrt* and *Pfdhps* are the result of the SP and AQ dynamics as administered in SMC will require *in vitro* and *in vivo* phenotypic validation. The hypotheses raised in this study will also need to be recontextualized with comprehensive models of drug resistance that consider how drug resistance mutations change in populations with multiple drug recommendations and regimens^37,38^. This is particularly relevant for interpreting the changes in *Pfmdr1,* where a variety of mutations and copy number variations have been shown to modulate resistance against multiple drugs^2,18^. While the *Pfmdr1* NFD haplotype has been used as a surrogate marker for AQ and lumefantrine resistance^39,40^, the observed changes in haplotype frequency likely reflect the combined impact of multiple drugs (**Fig 2D**). Dissecting the factors driving the changes in *Pfmdr1* and their implications for future therapeutic and chemopreventive strategies is a subject for future study.

Model-based assessments of mutation frequency would also be helpful for determining why post-2014 changes in *Pfdhps* A437G and *Pfcrt* K76T were observed in both SMC (Kedougou) and non-SMC regions (Thiès). One possibility is that the changes represent two separate selection events occurring in Thiès and Kedougou. The sharp increase in Thiès could be the result of unofficial CQ use or use of CQ for COVID-19 prevention or treatment in 2020^41^. However, the rise in *Pfcrt* K76T predates COVID-19, and we are unaware of any factors that might have caused a major shift in unofficial CQ use in 2014. Alternatively, this could indicate that the parasite populations in Senegal are interconnected and that parasites from SMC regions are being exported to non-SMC regions. Connectivity between Kedougou and Thiès is supported by the appearance of the major post-2014 *Pfcrt* K76T haplotypes at both sampling sites. Reports from the 2014 Senegal census show significant levels of human movement from the higher transmission, southeastern corner of the country (in which Kedougou is located) to the lower transmission western sections of the country (in which Thiès is located)^42^. Incorporating travel history surveys into molecular surveillance may help determine to what extent parasite populations are interconnected.

A major limitation of our study was that sample collection was uneven across space and time. To summarize the data, we combined the data into a single, Senegal-wide model based on the drug resistance marker data from Pikine, Thiès, and Kedougou. This approach assumes each study site represents a sampling from a greater, well-mixed parasite population. When examined separately, the data from each site followed the same temporal trends. In this study, evidence of a single, admixing parasite population included 1) the fact that Pikine and Thiès are geographically proximal (∼30 miles), 2) the post-2014 mutation frequencies in Thiès and Kedougou follow similar trajectories, and 3) the mutation frequencies observed in sites not included in the model (Kolda, and Kaolack) tended to be consistent with the model predictions. Previous parasite population genetic analyses in Senegal based on the time-serial allele frequency data of neutral sites and Fst from *msp*-typing have consistently suggested a well-mixed parasite population^43,44^. While we cannot exclude the possibility of site-specific differences in mutation frequency, there is little evidence that the parasite population genetic structure in Senegal is as fragmented as those seen in the Greater Mekong region of Southeast Asia^45^.

Likewise, our limited surveillance of the *Pfkelch13* limits our conclusions regarding artemisinin resistance in Senegal. Although we did not detect significant levels of either *Pfkelch13* C580Y or *Pfkelch13* A578S in Senegal, we did not examine the other mutations associated with delayed clearance in the *Pfkelch13* propeller domain in our SNP-based molecular surveillance^46–48^ or the *Pfkelch13* C439Y and A675Y mutations observed in Uganda^49–51^. Expanding molecular surveillance to include amplicon-based^52^ approaches to genotype the entire Pfkelch13 propeller domain would allow for better assessments of growing artemisinin resistance risk in Senegal.

To summarize, our molecular surveillance suggests widespread pyrimethamine resistance in Senegal and shows signs of emerging AQ resistance after the introduction of SMC due to an increase in *Pfcrt K76T*. Unusually, we detected a recent decrease in *Pfdhps* A437G, which could indicate a return to sulfadoxine susceptibility, which we hypothesized is due to a yet uncharacterized fitness cost when AQ is present. Given the increased drug pressure associated with using the same drug for both treatment and prevention, we recommend that AQ not be used as a partner drug in artemisinin combination therapies in SMC-treated regions (consistent with WHO recommendations) and that the chemopreventive efficacy of SMC be closely monitored^53^.

## METHODS

### Sampling

Parasite samples were collected from treatment seeking patients aged 3 months or older presenting with fever or history of fever within the past 48 hours in Pikine, Thiès, Kedougou, Diourbel, Kolda, and Kaolack between 2000 and 2020. Informed consent was administered (to parents or guardians if the patient was a minor). All patients with positive tests received free malaria treatment with AL or ASAQ, in accordance with the National Health Development Policy in Senegal as recommended by WHO.

In addition to slide preparation and RDT, all consenting patients also gave a finger stick blood sample spotted on filter paper (Whatman Protein Saver FTA-about five drops). After drying, samples were stored in plastic bags at room temperature and protected with silica gel desiccant for later DNA isolation for molecular testing.

### Laboratory procedures

#### Sample collection and DNA extraction

A total of 3,284 DNA samples were extracted from blood spots collected on Whatman Protein Saver FTA-filter papers (Whatman® 3MM CHR CAT N° 3030-662) using QIAamp DNA Mini Kit (QIAGEN, Valencia, CA, USA), according to manufacturer’s directions.

### High Resolution Melting

Drug resistance markers in *Pfcrt*, *Pfmdr1*, *Pfdhfr*, *Pfdhps* were assessed using High Resolution Melting (HRM) assay with the Roche LightCycler 96 instrument (Roche Molecular systems) as previously described^54^. The HRM assay was set up in a total volume of 5 μl containing 2.5 μl of DNA and 2.5x LightScanner mastermix LCGreen (Plus double-stranded DNA dye (Idaho Technology, Inc.)). The codons 76 in *pfcrt* gene; 86, 184, 1042, 1246 in *pfmdr1* gene; 51, 59, 108 in *pfdhfr* gene; 437, 540, 581, 613 in *pfdhps* gene were used. The following reference DNA strains were used: 3D7 (*Pfmdr1 NYD*; *Pfcrt* K76; *Pfdhfr* N51, C59, S108; *Pfdhps* A437, K540, A581, A613; *Pfkelch13* C580), Dd2 (*Pfmdr1* YFD; *Pfcrt* K76T; *Pfdhfr* N51I, C59R, S108N; *Pfdhps* A437G) HB3 (*Pfmdr1 NYD*; *Pfcrt* K76; *Pfdhfr* N51, C59, S108N; *Pfdhps* A437G, K540, A581, A613) Tm90C6B (*Pfmdr1 NYD*; *Pfcrt* K76; *Pfdhfr* N51, C59R *; Pfdhps* A437, K540, A581G*; Pfmdr 1 NYD*) and MRA1236 (*Pfkelch13* C580Y).

### DNA Sequencing

The *Pfkelch13* propeller domain was amplified using a *P. falciparum* specific protocol described previously^55^. PCR products were visualized on a 2% agarose gel after electrophoresis. Sequencing of PCR products was performed using an ABI 3730 sequencer by Sanger using a protocol established at CDC/Atlanta Malaria Genomic laboratory (Applied Biosystems, Foster City, CA).

### Data Analysis

The HRM result was analyzed using the LightCycler 96 application software version 1.1.0.1320. The *Pfkelch13* sequence data was analyzed using the Geneious software (www.geneious.com). A cutoff of quality score HQ>30% was applied to all sequences. Polymorphisms were considered if both the forward and reverse strands carried a mutation and matched the quality score cut off.

### Haplotype Analyses

A subset of 231 monogenomic (single-strain) samples collected from febrile, clinic-reporting patients between 2006 and 2019 from Pikine, Thiès, and Kedougou were previously whole genome sequenced using next-generation Illumina short reads. These sequences excluded polygenomic (multiple strain) infections and avoided repeated sequencing of clones using a 24-SNP molecular barcode.

Briefly, short-reads were aligned to the *P. falciparum* 3D7 reference genome (PlasmoDB v. 28) using BWA-mem and Picard Tools. Variants were called using HaplotypeCaller in GATK v3.5. Our whole genome sequence analyses focused on a set of 577,487 SNPs spread throughout the core region of the genome. Bifurcation plots and extended haplotypes were calculated using *rehh 2.0*^56^.

These whole genome sequences were used to examine the genomic region surrounding *Pfcrt, Pfdhfr,* and *Pfdhps*. Genomic regions were defined as the region 100kb upstream and downstream of the starting and ending boundaries of each gene. The gene boundaries for *Pfcrt, Pfdhfr,* and *Pfdhps* were obtained from Plasmodb (Plasmodb.org).

For each genomic region surrounding *Pfcrt, Pfdhfr,* and *Pfdhps*, samples and variant sites were filtered by 1) excluding samples with > 50% unusable data in the examined genomic region, 2) retaining sites with < 10% unusable data in the remaining samples, and 3) further excluding samples with > 5% unusable data in the retained sites. Unusable data was defined as any site with missing data (failed to be genotyped), or a site that was heterozygous or triallelic. These filters were applied separately across all sample years, for samples collected before 2014 (pre-SMC), and for samples collected after 2014 (post-SMC).

When applied across all sample years, the *Pfcrt* genomic region included 171 samples and 173 SNPs, the *Pfdhfr* genomic region included 170 samples and 85 SNPs, and the *Pfdhps* genomic region included 182 samples and 83 SNPs. When applied on the pre-SMC data, the *Pfcrt* genomic region included 64 samples and 21 SNPs, the *Pfdhfr* genomic region included 54 samples and 27 SNPs, and the *Pfdhps* genomic region included 51 samples and 75 SNPs. The majority of these samples were collected after 2010. For the post-SMC samples, the *Pfcrt* genomic region included 114 samples and 213 SNPs, the *Pfdhfr* genomic region included 170 samples and 95 SNPs, and the *Pfdhps* genomic region included 127 samples and 89 SNPs (**Fig S5**).

### Generalized Additive Model

Binomial generalized additive models (GAMs) were fit using the R package *mcgv* (v1.8-40). The GAM quantifies the relationship between mutation frequency and sampling year across all sites, while considering sample size and sampling origin. The structure of the GAM was defined as:

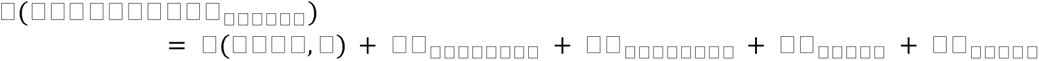

where 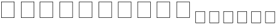 is the proportion of samples with the mutant allele, *s*(*year, k*) is a smoothing spline function for the sample year. The *k* is the degrees of freedom used in the smoothing spline function. The other covariates are binary categorical variables specifying sample origin. Kolda and Kaolack were not included in the model because they were sampled for less than three years.

### H12 and H2/H1 Analyses

Genotypes were called for sequenced samples at 149,582 SNP sites, sites that were chosen as reliable based on the full Pf3k dataset; details of the filtering are described in another manuscript^57^. For this analysis, an additional 10,387 SNPs were eliminated because of incompatible allele calls for 2019 and 2020 data (although only the former are being reported here). These were then further filtered to remove sites that were monomorphic or rare in our dataset, as well as filtered to remove densely packed sites. Specifically, a threshold on the minimum minor allele frequency was set at 0.5%, while the minimum spacing between markers was 20 bp and the maximum number of SNPs allowed in a 2 kb window was 12. Within these restrictions, SNPs were prioritized by minor allele frequency. The remaining 43,846 SNPs were used for analysis.

Rather than calculating these statistics directly from sequence data, however, we instead based them on shared chromosome segments that were identified as being identical by descent. Haplotype sharing among parasites was determined by first identifying chromosome segments that were identical by descent (IBD) between each pair of parasites, using the program hmmIBD (version 2.0.4)^58^. Because in hmmIBD, the threshold detecting IBD segments between two parasites depends on their genome-wide relatedness, a modified version of the program was used which fixed the genome-wide IBD fraction at 20%. In this way, a determination of local IBD status could be made without bias by the parasites’ overall relatedness. We adopted this IBD approach because the hidden Markov model that it is based on is well suited to identifying closely related haplotypes in our dataset. It takes into account differing SNP frequencies, does not rely on arbitrary window sizes, and incorporates into the model sequencing error and missing data.

At each SNP locus, haplotypes were defined by clustering samples that were related to one another. Clustering was performed via a greedy algorithm as follows. For a pair of samples IBD at a SNP site:

⍰ If neither is already in a cluster, form a new cluster.
⍰ If both are already in the same cluster, do nothing.
⍰ If they are already in different clusters, merge the clusters if 40% of pairwise comparisons are IBD.
⍰ If one is in a cluster, add the other if it is IBD with at least 40% of existing samples in the cluster. Otherwise, start a new cluster.

The resulting clusters are defined as the set of haplotypes at that locus. If we label the population frequency of these haplotypes as p_1_, p_2_, p_3_, etc., we then calculated H1, H2, and H12^20,59^:

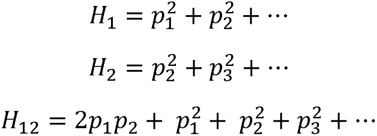

H12 treats the two most common haplotypes as a single haplotype. In these calculations, singleton haplotypes are omitted since their population frequency is likely to be much lower than their sample frequency.

### Ethics statement

The study was reviewed and approved by the Ministry of Health and Social Action in Senegal (Protocol SEN14/49) and the Institutional Review Board at Harvard TH Chan School of Public Health (CR-16330-07). This study was registered at the Pan African Clinical Trials Registry on 09 March 2020 under the number PACTR 202003802011316. The work was supported by the NIH/ICEMR, International Centre of Excellence for Malaria Research, west Africa (U19AI089696) and the Bill and Melinda Gates Foundation (0PP1200177).

## Data Availability

All data produced in the present study are available upon reasonable request to the authors

## Supplemental Figures

**Fig S1.**
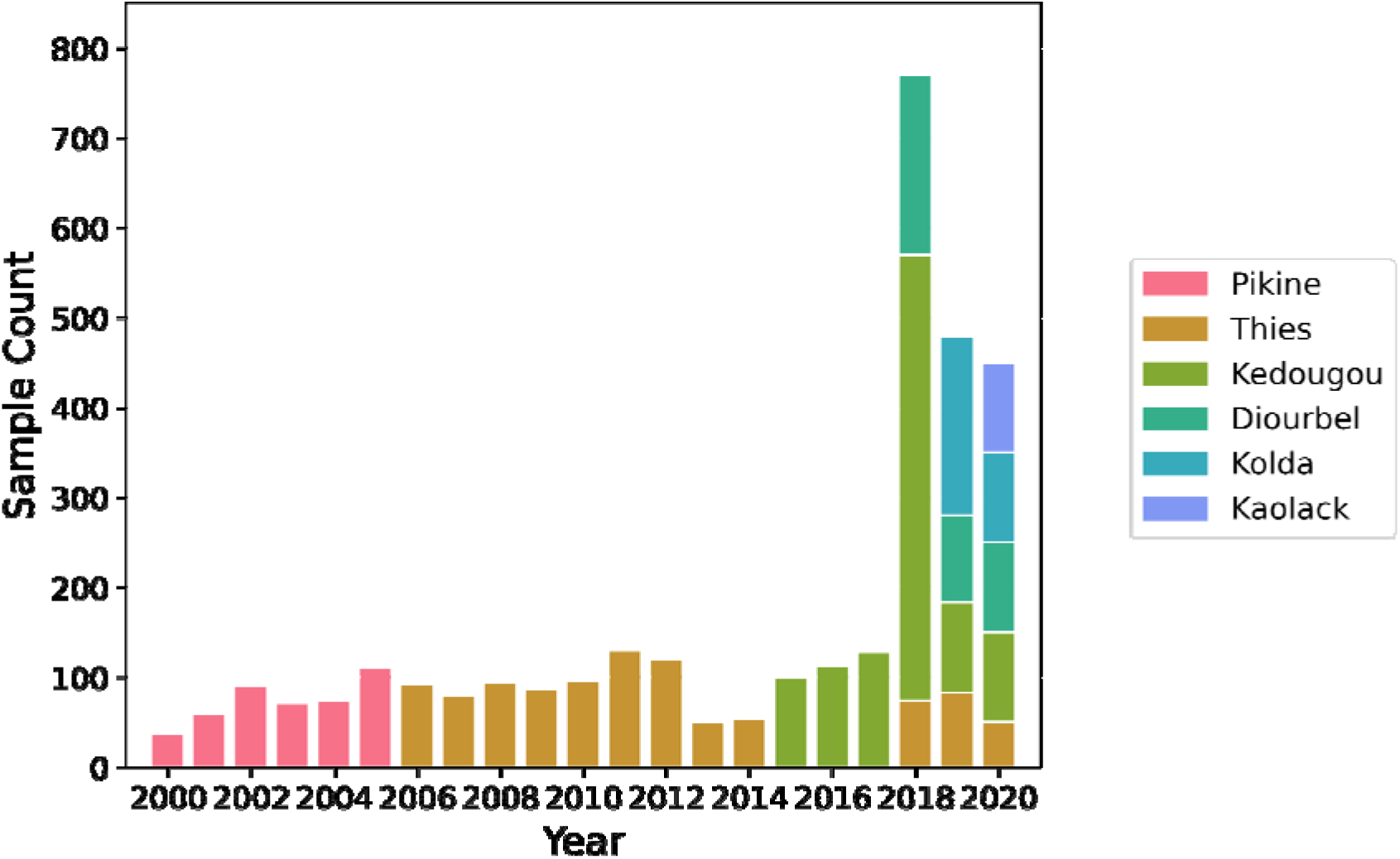
Sample sizes for SNP-based molecular surveillance. Sample size per year per region for the SNP-based molecular surveillance.

**Fig S2.**
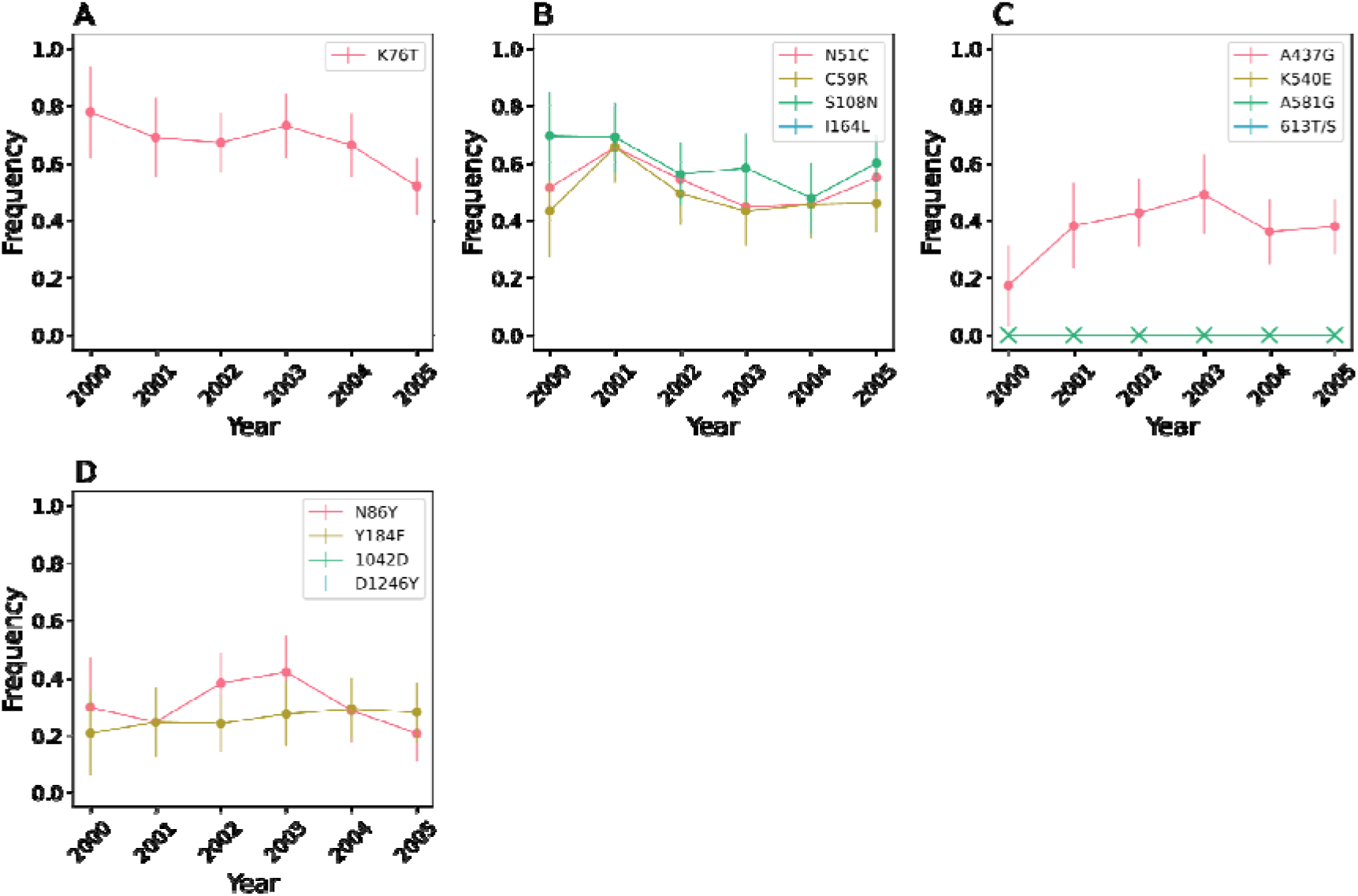
SNP-based molecular surveillance in Pikine. **A**) Pfcrt, **B**) *Pfdhfr*, **C**) *Pfdhps* and Pfmdr1. The *Pfkelch13* SNPs were not examined in Pikine. Error bars indicate two binomial standard deviations from the mean. X’s denote years where samples were collected but the mutation was not observed. Gaps in the data were because samples were not collected for that year.

**Fig S3.**
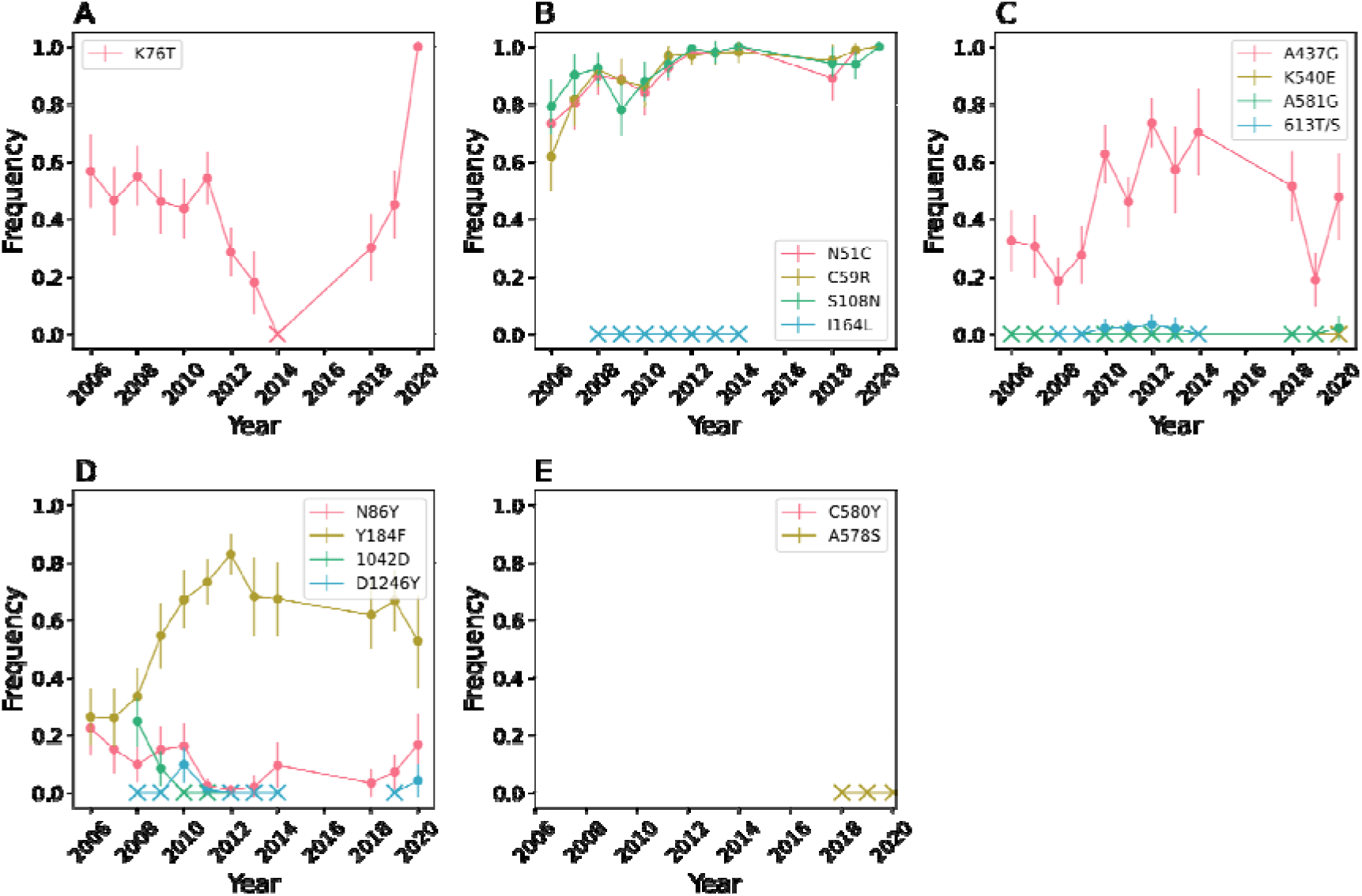
SNP-based molecular surveillance in Thies. **A**) Pfcrt, **B**) *Pfdhfr*, **C**) *Pfdhps* and **D**) Pfmdr1, and **E**) *Pfkelch13*. Error bars indicate two binomial standard deviations from the mean. X’s denote years where samples were collected but the mutation was not observed. Gaps in the data were because samples were not collected for that year.

**Fig S4.**
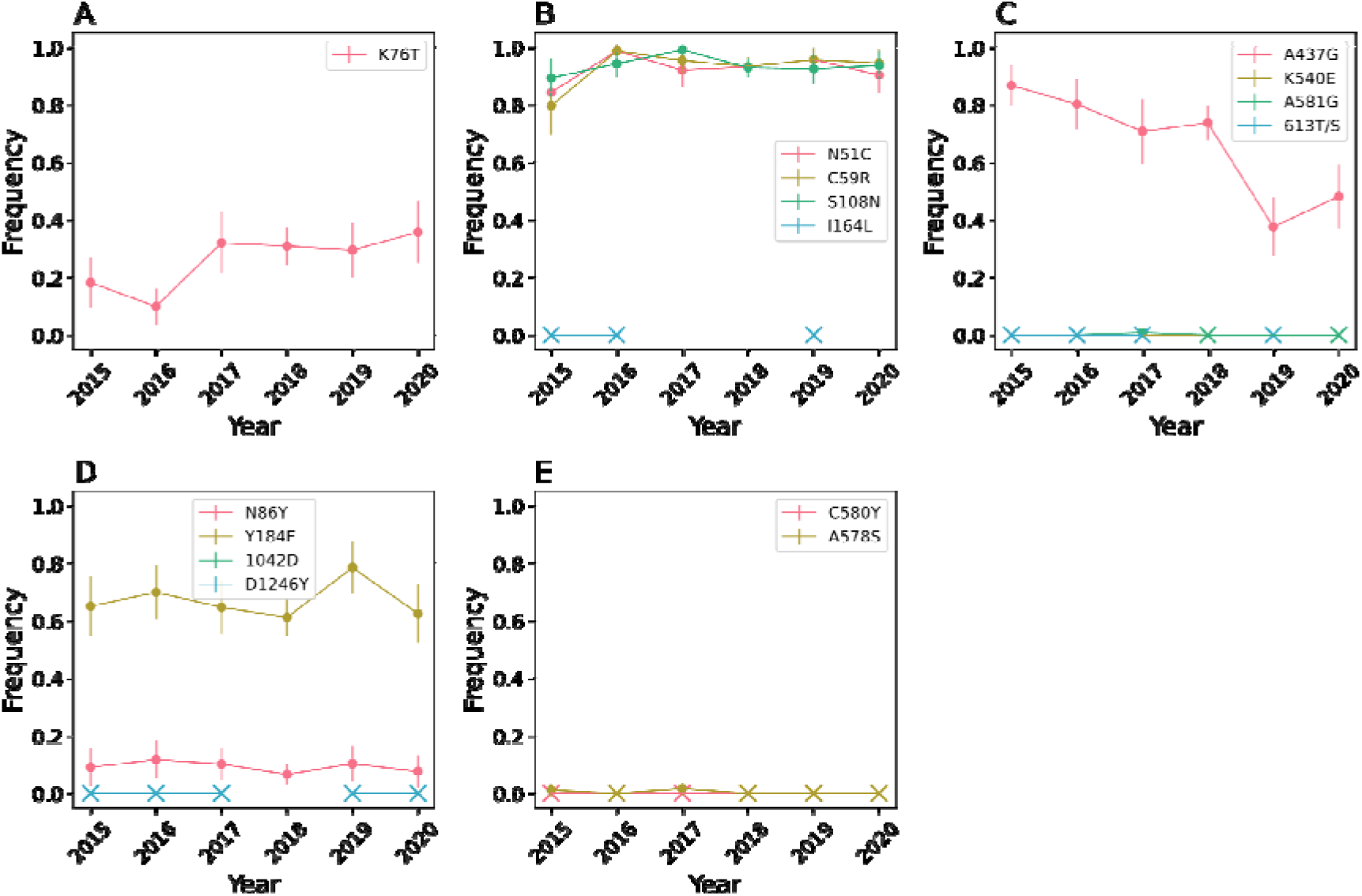
SNP-based molecular surveillance in Kedougou. **A**) Pfcrt, **B**) *Pfdhfr*, **C**) *Pfdhps*, **D**) Pfmdr1, and **E**) *Pfkelch13*. Error bars indicate two binomial standard deviations from the mean. X’s denote years where samples were collected but the mutation was not observed. Gaps in the data were because samples were either not collected or not genotyped for that year.

**Fig S5.**
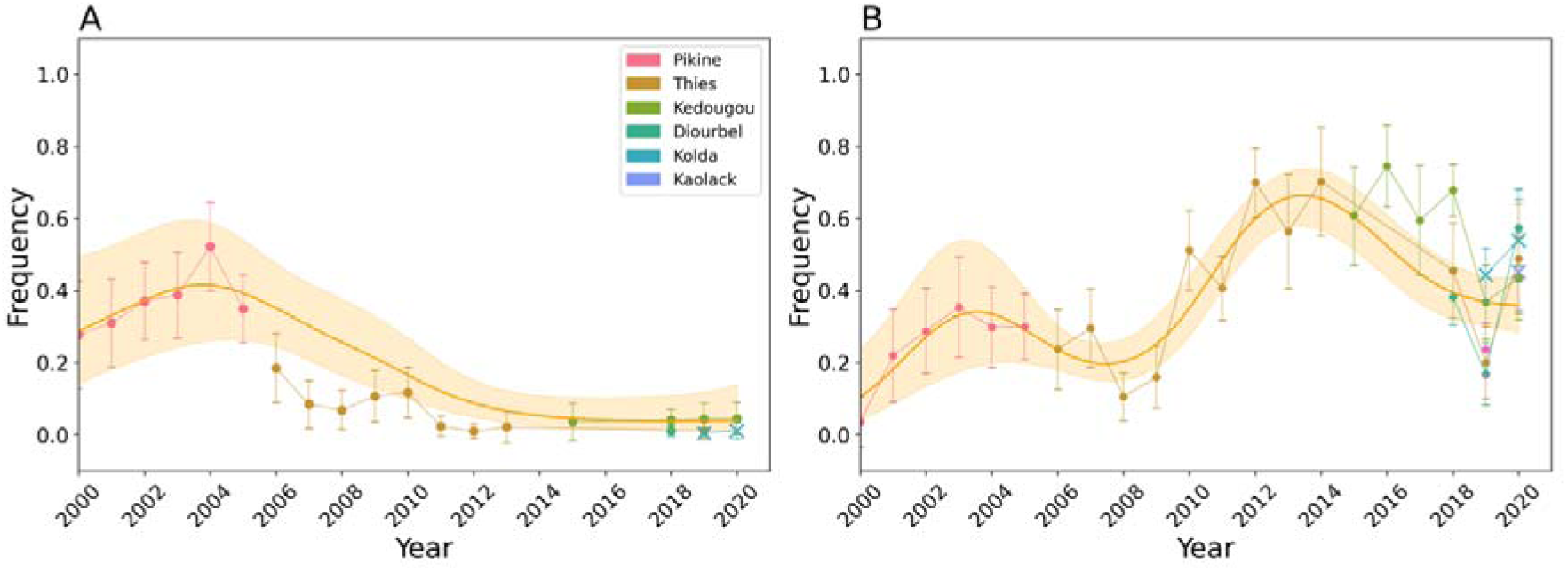
**A**) Frequency of *Pfdhfr* triple sensitive (N51, C59, S108) parasites. **B**) Frequency of “quadruple” (*Pfdhfr* triple mutant + *Pfdhps* A437G) parasites. The scatterplots show the observed frequencies and their 95% binomial confidence interval. Model predictions from a calibrated generalized additive model and the 95% confidence intervals are shown in orange. The model was calibrated with data from Pikine, Thiès, Diourbel, and Kedougou (denoted with circles). The data from Kolda and Kaolack (denoted with X) were not used for model calibration.

**Fig S6.**
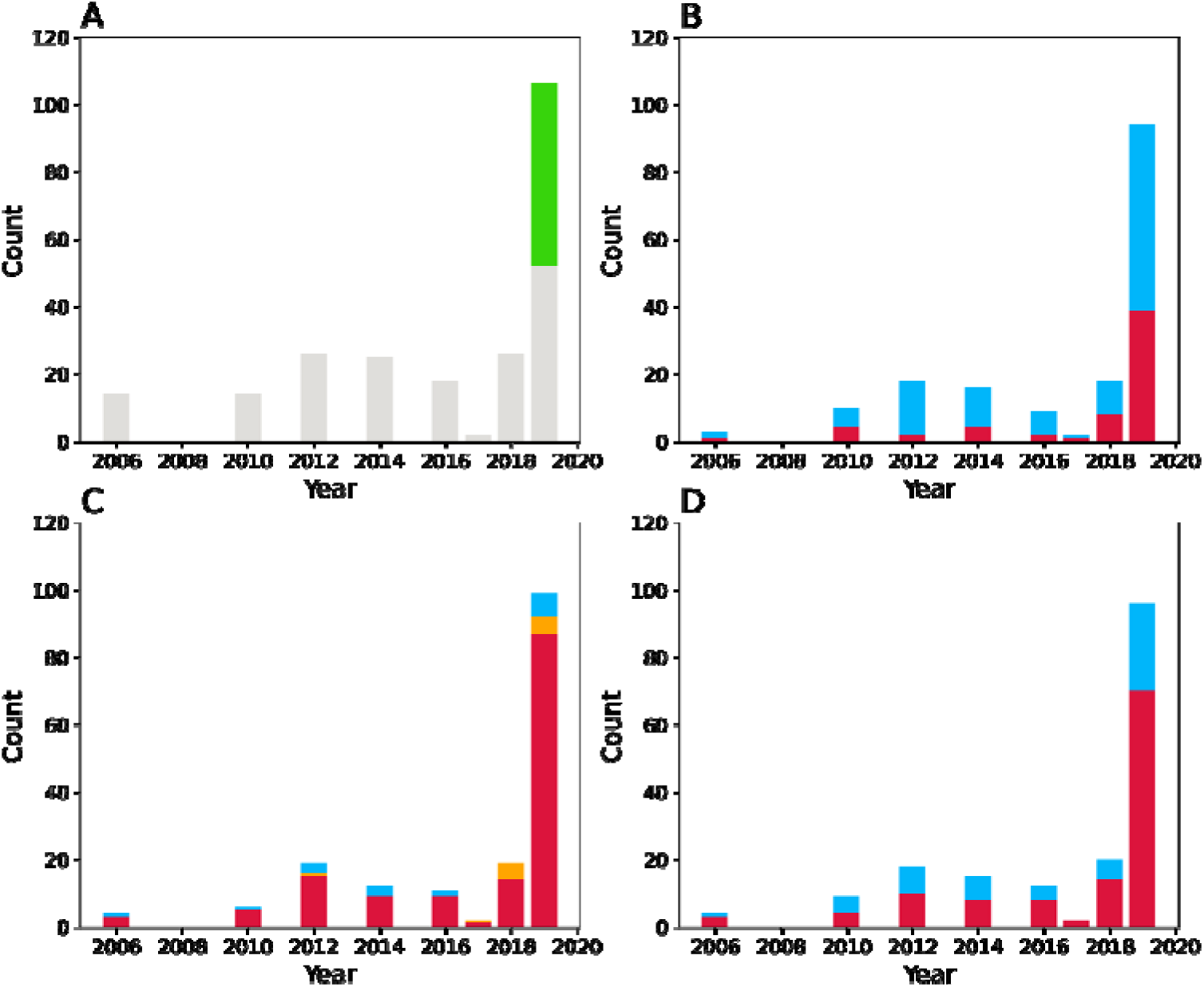
Sampling distribution for our whole genome sequence collection. (**A**). Grey indicates the sample came from Thiès. Green indicates the sample came from Kedougou. Sampling distributions for **B**) the *Pfcrt* genomic region, **C**) the *Pfdhfr* genomic region, and **D**) the *Pfdhps* genomic regions. For **B** and **D**, blue denotes samples with the sensitive allele and red indicates those with the resistance allele. For **C**, red denotes samples that are *Pfdhfr* triple mutant, blue indicates those that are *Pfdhfr* triple sensitive, and orange indicates those with a mix of resistant and sensitive alleles at the three examined *Pfdhfr* loci.

**Fig S7.**
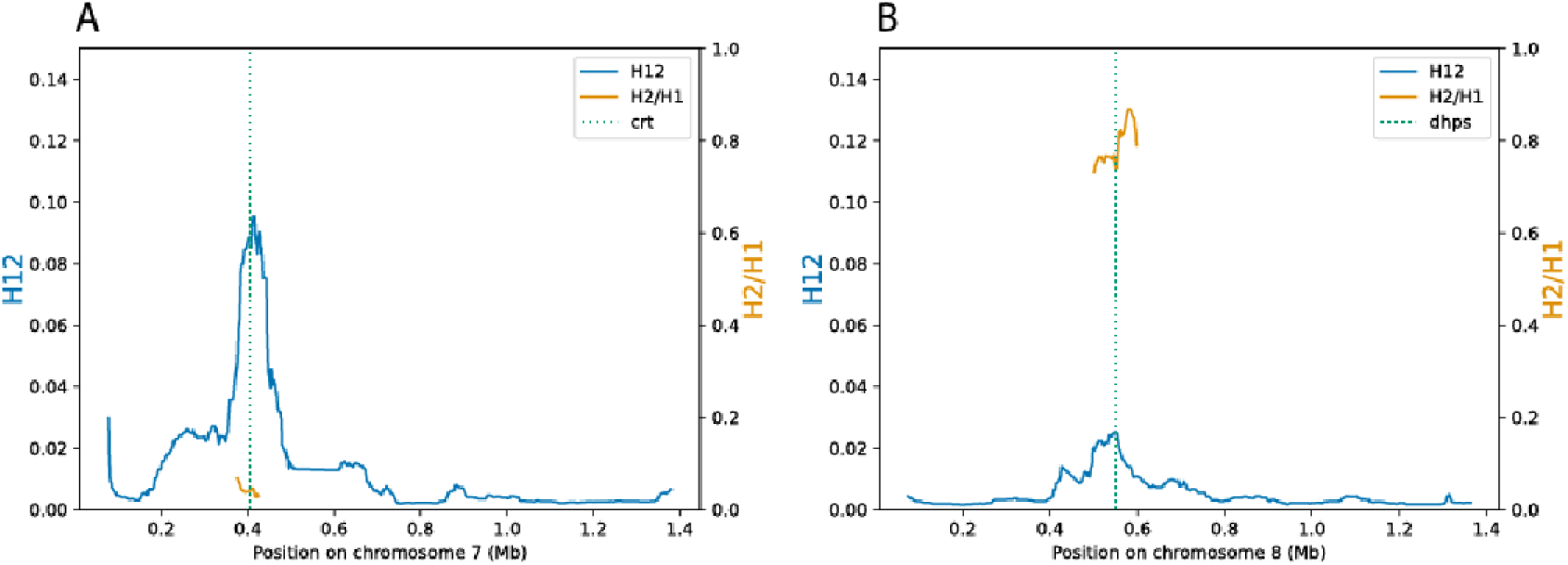
Evidence of Hard and Soft Sweeps. H12 (*blue*, left y-axis) and H2/H1 (*orange*, right y-axis) statistics for **A**) chromosome 7 and **B**) chromosome 8. The dotted green lines show the location of *Pfcrt* or *Pfdhps*.

**Fig S8.**
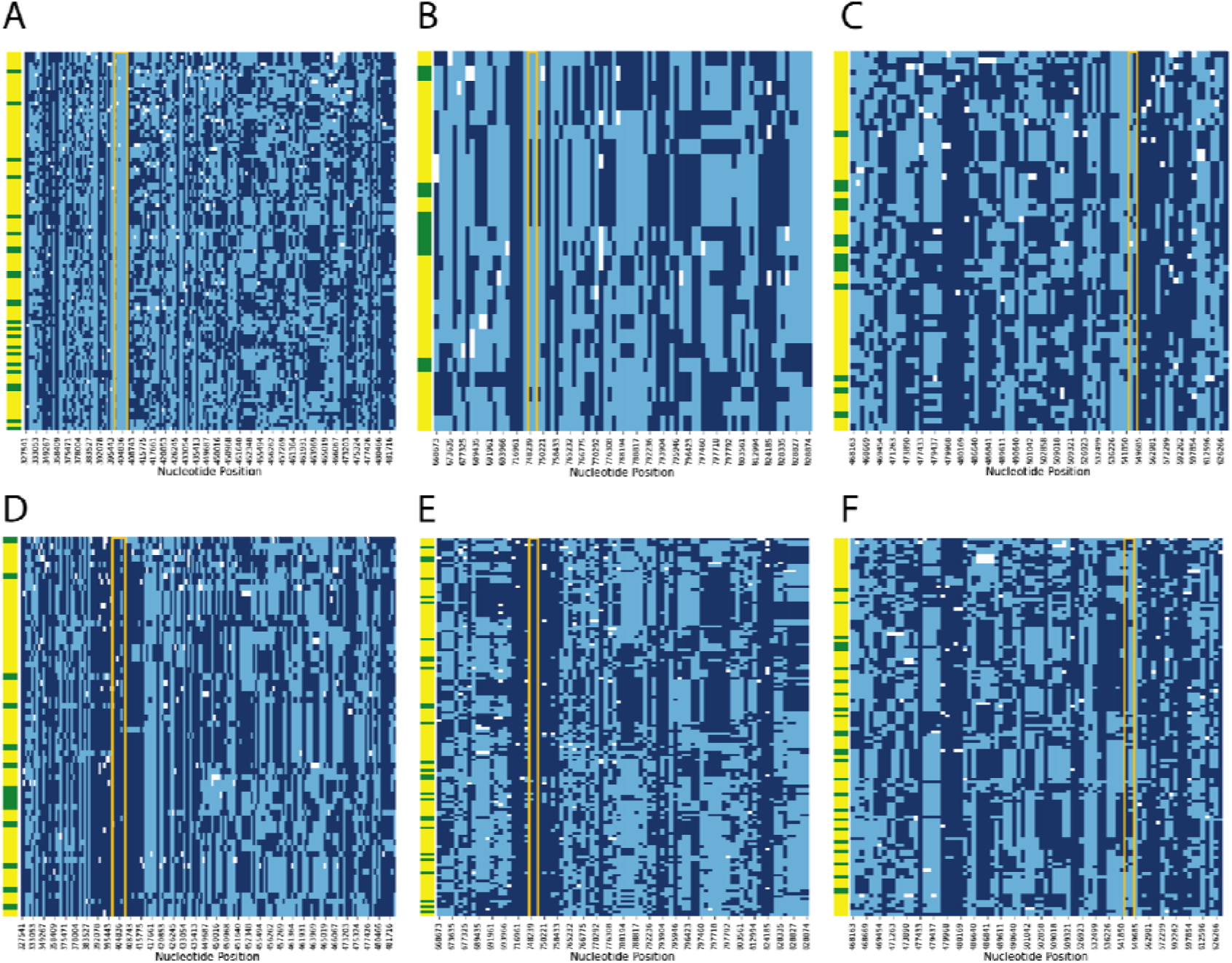
SNPs used to define genomic haplotypes. Genomic haplotypes surrounding the wild-type mutations: **A**) *Pfcrt* K76, **B**) *Pfdhfr* C59, **C**) *Pfdhps* A437 and the drug resistance mutations: **D**) *Pfcrt* K76T, **E**) *Pfdhfr* C59R, **F**) *Pfdhps* A437G. Each row represents a sample. The left most column indicates whether the sample was collected before 2014 (*green*) or after 2014 (*yellow*). Alleles corresponding to the 3D7 reference are indicated by *light blue* and alleles corresponding to the alternative allele are indicated by *dark blue*. White corresponds to missing data. The orange boxes highlight the boundaries of the *Pfcrt* (**A/D**), *Pfdhfr* (**B/E**), and *Pfdhps* (**C/F**) genes,

